# Non-obese population with the rs7903146 T allele exhibits higher sugar and more dyslipidaemia in subjects with Type-2-diabetes

**DOI:** 10.1101/2020.09.09.20186791

**Authors:** Sarah Shaibu, Ishaya Yohanna Longdet, Carrol Domkat Luka, Jesse Fanen Ortswen, Gloria Eleọjọ Eneọjọ-Abah, Joel Iko-Ojo Oguche, Tijani Salami, Shedrack Egbunu Akor, Samuel Eneọjọ Abah

## Abstract

Type 2 diabetes (T2D), the most prevalent type of diabetes has been associated with Transcription-Factor-7-Like-2 gene Single Nucleotide Polymorphisms (SNPs), rs12255372 and rs7903146 as risk factors, thought to be modulated by obesity status. In sub-Saharan Africa, the onset of T2D in the non-obese is rarely suspected. This study looks into the genetics and the biochemical parameters in non-obese population, with and without T2D and living in Jos, Nigeria. A total of 68 subjects, 40 diabetic patients and 28 healthy control group, all with closely matched age, height, nutrition, family history, Body Mass Index and socioeconomic status, recruited from within the same population were studied. SNPs Genotyping were performed using Polymerase Chain Reaction and Sangers Sequencing. Lipid profiles, Fasting Blood Sugar and C-peptide levels were measured and analysed alongside with demographic data from questionnaire. Odd-ratio at 95% confidence interval at a conventional level of alpha, <0.05 and Product Moment Correlation Coefficient Analysis were used to analyse the data in both groups. The entire population showed the GG genotype for the rs12255372. However, different genotype combination, CC, CT and TT were observed with the rs7903146. Though no significant association was observed between the genotypes and the odd of T2D, healthy subjects with the T allele showed a higher level of two hours postprandial plasma glucose level than those with CC genotype. Patients with T allele shows a more abnormal level of diabetes metabolic syndrome indicators such as Fasting Blood Sugar; two hours postprandial plasma glucose level; C-peptide; Low Density Lipoprotein, High Density Lipoprotein and Total Cholesterol. The study suggests that lower sugar metabolism and more dyslipidaemia are observed in subject with T allele. Hence, this could constitute poorer prognosis and a risk factor for non-obese population, particularly with high carbohydrate intake.

## INTRODUCTION

Diabetes is a chronic metabolic disorder characterized with an abnormally high sugar level in the blood and are of different types, type-1-diabetes; type-2-diabetes; gestational diabetes and autoimmune diabetes(1, 2). According to World Health Organisation (WHO), Type-2-Diabetes (T2D) accounts for about 90% of the diabetes around the world. Two out of every three persons remained undiagnosed in the sub-Saharan Africa. In Nigeria, the 2016 WHO country report indicated about 73, 000 diabetes associated deaths of which 58.4% were women.

Type-2-diabetes (T2D) cases results from the interaction between genetic and environmental factors (3–7). Genome Wide Association Study (GWAS) has been used to identify a number of genetic marker associated with diabetes (8). The strongest and widely reported marker across populations is the rs7903146, which is a Single Nucleotide Polymorphism (SNP) on Transcription Factor-7-Like-2 (TCF7L2) gene on chromosome 10 (8–10).

The rs7903146 is reported to play a role in both α and β pancreatic cell metabolism and proglucagon gene expression in intestinal enteroendocrine cells(11, 12). The T allele has been reported to show association with impaired insulin secretion, incretin effect and enhanced rate of hepatic glucose production (13). Each risk allele is thought to increase the risk of T2D by 1.34 times, making the TT genotype the strongest predictor of T2D (9, 13,14). However, these reports are not only conflicting across populations but that the risk are reported to be modulated by obesity status. Within the European population, the rs7903146 genotypes is thought to confer a more significant risk of T2D in non-obese subjects, BMI < 30kg/m^3^, (15, 16). This SNP was also thought to have been associated with protection from obesity (17) and lower BMI (18).

Obesity is widely claimed to predispose to diabetes (19–21). Unlike the obese group that are constantly being advised on nutrition and exercise, the onset of type-2-diabetes (T2D) in the non-obese group is rarely suspected. A plausible genetic risk factors among this group is rs7903146 genotypes (7, 23,16, 17). We therefore hypothesize that the rs7903146 genotypes is associated with increased risk of T2D and a higher T2D metabolic syndromes indicators in the non-obese population in Jos, Nigeria.

Since rs7903146 SNP appear to exhibit marked environmental variation owing to marked differences across population, it is important to investigate the role of this SNP among the no-obese subject in sub-Saharan Africa. This paper tends to answer whether or not the rs7903146 genotypes are associated with T2D among the non-obese group in Jos, Nigeria and whether or not the genotypes are important in the management of T2D in the non-obese population.

## MATERIALS AND METHODS

### Ethical Approval, Study Population and Study Site

This research was approved by University of Jos, Nigeria and the EBIHAS Research Group. Consents were obtained from study participants in accordance with the World Medical Association Helsinki declaration statement of ethical principles for medical research involving human subjects.

A total of 68 non-obese subjects were recruited, 40 patients and 28 controls (Table 1). The patient group were recruited at Faith Alive Hospital, Jos, Nigeria, where they were also receiving treatments. The control group were apparently healthy subjects recruited from outpatient clinic and home visit, within the same population in Jos, Nigeria. The population studied were unrelated individuals, but with similar sociodemographic factors. Jos is located on the plateau, about 3800–4100ft above sea level. It’s a region in Nigeria with temperature as low as 7–11°C in Harmattan (a season in West African Subcontinent, which occur between November and Mid-March and characterized with cold, dry and dusty north-easterly wind trade from the Sahara Desert) and up to 11–25°C on the average all through the year.

**Table 1:** The Cohort Table.

| Variable | Patients | Control | P-value |
| --- | --- | --- | --- |
| Number | 40 | 28 | na |
| Age(Median IQR) | 27.5 (21-43) | 27 (20-42) | ns |
| Sex (M/F) | 19/21 | 14/14 | na |
| Height (m<sup>2</sup>; Median IQR) | 1.7 (1.62-1.77) | 1.6 (1.57-1.73) | ns |
| Weight (kg; Median IQR) | 71(61.8-75.3) | 69 (62-73) | ns |
| Alcohol Consumption (%) | 43.00% | 43.45% | na |
| Smoking (%) | 15.50% | 12.00% | na |
| Family History (%) | 25.00% | 14.00% | na |
| Blood pressure (mean) | 142/80 | 131/78 | na |
| Nutrition (CHOs), Average | 42.50% | 41.00% | ns |
| BMI, kg/m<sup>3</sup>(Median IQR) | 23.9 (22.3-26.3) | 23 (21-24) | ns |

IQR = Interquartile range; ns = Not Significant; na = Not Applicable; 2-h = Two (2) hour; CHOs = Carbohydrates; BMI = Body Mass Index; FBS = Fasting Blood Sugar; M/F = Male/Female. The frequency of alcohol consumption, nutrition and smoking per week were expressed in percentage.

### Case Definition and Diagnosis

The patient group consists of individuals who were diagnosed with diabetes according to the World Health Organization (WHO) criteria, fasting plasma glucose ≥ 7.0mmol/l (126mg/dl) or two hours postprandial plasma glucose ≥ 11.1mmol/l (200mg/dl). The control group consists of healthy non-diabetic subjects considered to have normal plasma glucose level in accordance to WHO criteria, fasting plasma glucose < 6.1mmol/l or two hours postprandial plasma glucose < 7.8mmol/l. Co-founding factors like drugs in the family of sulfonylureas were yet to be administered to patients prior to the test.

### Data Collection

We used a tailored questionnaire to collect both data and blood samples. The study population consisted of both male and female with similar demographic data (Table 1) and closely matched variables like alcohol consumption, smoking, Blood pressure, nutrition and family history (Table 1). Blood samples were collected and assayed for total cholesterol, triglycerides (TGs), High density lipoprotein (HDL), low-density lipoprotein (LDL), sugar and C-peptide. DNA were extracted from the blood for genotyping. The rate of carbohydrate and alcohol consumption were determined from the questionnaire based on the number of consumptions per week.

### Oral Glucose Tolerance Test (OGTT) – Two (2) Hours postprandial Plasma Glucose

Prior to the OGTT, the study groups were advised to fast except water and without smoke and coffee and activity for two (2) hours. An initial venesection was done prior to administration of glucose solutions and was considered as initial values. A 75g glucose solution was administered to each subject prior to venesection, 2 hours after consumption.

### Measurement of Serum Glucose

Whole blood was collected into the fluoride oxalate bottle and allowed to settle by leaving it undisturbed at room temperature for 15–30 minutes. The plasma was separated from the whole blood by centrifuging at 1, 000–2, 000 x g for 10 minutes in a refrigerated centrifuge and the resulting plasma was used. The determination of plasma glucose was based on enzymatic oxidation in the presence of glucose oxidase (equation 1) and peroxidase (equation 2). The hydrogen-peroxide formed (equation 1), reacts under catalysis of peroxidase with phenol and 4-aminophenozone (4-aminoantipyrine) to form a red coloured dye, quinoneimine (equation 1 and 2).

Briefly, test-tubes were labelled as Test, Standard and Blank respectively and to each of these three tubes, 1mL of the reagent (Phosphate buffer, pH7, 0.1mol/l; phenol, 11.0mmol/l; glucose oxidase, 1.5KU/l; peroxidae, 2KU/l and 4-aminoantipyrine, 0.4mmol/l) was added. To the blank, standard, and test specimen were also added 10µl of distilled water; glucose standard and serum respectively and were mixed and incubated for 10 minutes at 37°C. The absorbance of test and standard were read against the blank within 60 minutes at 546nm and the serum glucose concentration measured by dividing the absorbance of the test-specimen by that of standard and multiplied by a standard conversion value. The result of this procedure was validated with ACCU-Chek glucose measurement kit, following manufacturer’s procedure.

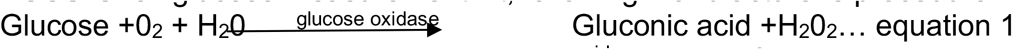

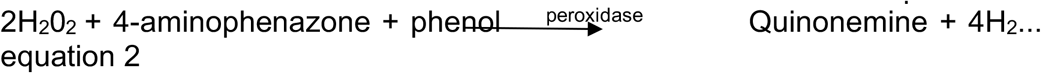

### Measurement of C-Peptide

The level of C-peptides were measured by ELISA. Briefly, a microplate was coated with biotin conjugated anti-human C-peptide monoclonal antibody (Invitrogen). Plate was incubated for an hour at room temperature with a coating buffer. After washing and blocking steps, an appropriate serum dilution was added and incubated, followed by another washing steps prior to addition of streptavidin-HRP and tetramethyl-benzidine (TMB). A 2M tetraoxosulfate-VI-acid solution was used as the stop solution and plate was read within 5 minutes at 490nm using a FLUorStar-Omega (BMG-Labtech) ELISA plate reader. The concentrations were computed using the five to four parameters logistic (5/4-PL) curve-fit (MasterPlex ReaderFit v2.0, MiraiBio Group, Hitachi Solutions America, Ltd).

### Determination of Triglycerides

The level of triglycerides in the serum was determined as previously described(25–27) with triglycerides kit (Randox) and based on glycerol-3-phosphate oxidase (GPO) and peroxidase (POD). The hydrolysis of triglycerides was catalysed by lipase to produce glycerol and free fatty acids (equation 3). The glycerol generated is then phosphorylated by adenosine 5’-triphosphate (ATP) in the presence of glycerol kinase (equation 4). Oxidation of the resulting glycerol 3-phosphate by GPO produces hydrogen peroxides (equation 5). Hydrogen peroxide in the presence of peroxidase (POD) reacts with 4-aminoantipyrine and 4-Chlorophenol to produce Quinonemine dye compound, as indicator (equation 3–6).

Briefly, 1 mL of enzymatic reagent was added to each test-tube labelled test-specimen, standard and blank. To the test-specimen, standard and blank were added 10µl of serum, glycerol standard and distilled sterilised water respectively and were mixed and incubated at 37°C for 10 minutes. Absorbance were measured at 546nm against the blank and the level of triglycerides was determined.

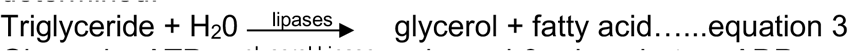

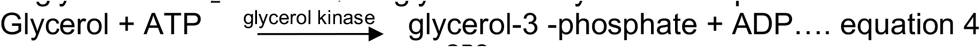

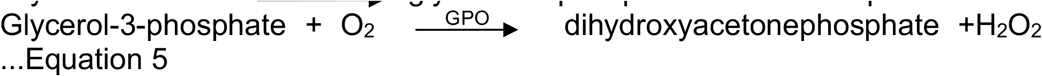

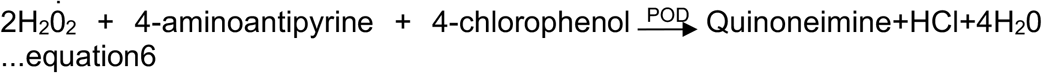

### Determination of Total Cholesterol

The levels of cholesterol were determined by a method of enzymatic hydrolysis as previously described (28, 29) and following the manufacturer’s instruction (Randox kits). The cholesterol is determined after enzymatic hydrolysis and oxidation: Cholesterol ester is hydrolysed to free cholesterol and fatty acids by cholesterol esterase (equation 7). Free cholesterol is then oxidized into cholesten-3-one and hydrogen peroxide in a reaction catalyzed by cholesterol oxidase (equation 8). The peroxidase catalyses the reaction between H_2_O_2_ and 4-amino phenazone and hydroxyl benzoate to produce quinoneimine (equation 9), a red coloured dye, whose absorbance was measured at 500nm after 5 minutes incubation at 37°C. The quinoneimine intensity is proportional to the total cholesterol present in the sample.

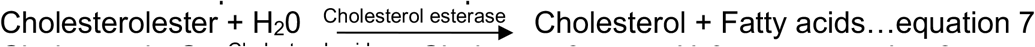

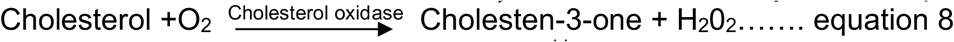

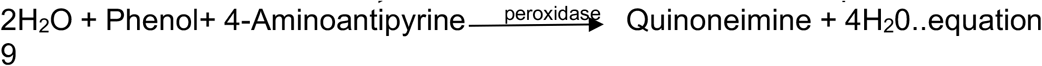

### Determination of HDL

High Density Lipoprotein (HDL) was determined as previously described (30) using HDL-cholesterol precipitant Kit (Randox). 0.5ml of serum was added to 1ml of HDL cholesterol precipitant, mixed and allowed to stand at room temperature for 10minutes and was centrifuged for 10minutes at 4000 rpm. The supernatant (0.1mL) was added to 1mL of CHOD-PAP to determine HDL cholesterol. The absorbances of the test-specimen and standard were measured against the blank within 60 minutes at 520nm after incubating for 5 minutes at 37°C.

### Low Density Lipoprotein (LDL) determination in mmol/L

LDL cholesterol = Total cholesterol – Triglycerides/2.2 – HDL Cholesterol.

### DNA extraction and PCR

Genomic DNA purification kit (GeneJET, ThermoFischers) was used for DNA extraction from blood following manufacturer’s instruction. The DNA was quantified with nanodrop (Thermoscientific).

Specific forward primers, 5’GGTGACAAATTCATGGGCTTTCTCTGCC3’ and reverse primer 5’AGAGATGAAATGTAGCAGTGAAGTG3’ spanning the region of interest (112, 998, 530–112, 998, 739; GRCh38.p12) and containing the SNP(rs7903146, at position 112, 998, 590; GRCh38.p12) was designed and used to extract and amplify a gene product size of 209bp by PCR with GoTaq master mix, 2x Promega, M7123 (12.5µl); both forward and reverse primers (1µl, 10µM each) and template DNA (2µl, 50ng), made up to 25µl total volume with 8.5µl nuclease free water. The PCR reaction was performed at 94°C for 5 minutes as initial denaturation; and 35 cycles of denaturation at 95°C for 30 seconds; annealing at 62.6°C for 30 seconds and extension at 72°C for 30 seconds. The final extension step was performed at 72°C for 5 minutes and thereafter held at 10°C at the end of the reaction. Electrophoresis was performed with 1% agarose; 1X TAE buffer (40mM Tris-acetate, 1mM EDTA, pH 8.3); SYBR safe DNA gel stain and hyper-ladder, 1kb (Bioline) and the right product size visualised and photographed using a gel documentation apparatus (Fig. 1A).

**Figure 1a:**
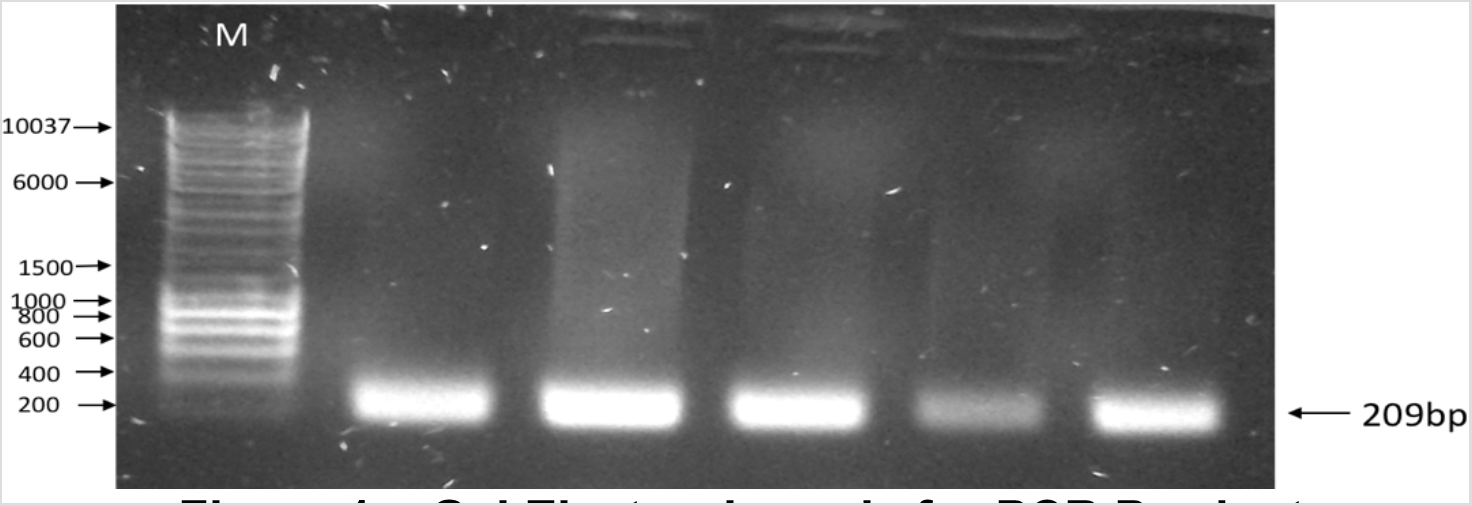
Gel Electrophoresis for PCR Product. Abbreviations: M = 1kb Molecular Hyper ladder (Bioline); bp = Base pairs. Numbers represent band sizes. The 209bp represent the right PCR product size from different samples that was sequenced.

### Sequencing and Genotyping

The individual PCR products were purified with a QIAquick PCR purification kit (QIAGEN) and quantified using Nanodrop (Thermoscientific). About 5µl (10ng/µl) of the product and forward primer (3.2pmol/µl) were sent to Source Bioscience for sequencing. Data were received in both SEQ and ABI formats. The ABI format was aligned with the reference sequence (ENSEMBL, GRCh38.p12) using SeqMan Pro alignment tool to identify the SNPs, (rs7903146) genotypes (Fig. 1B).

**Figure 1b:**
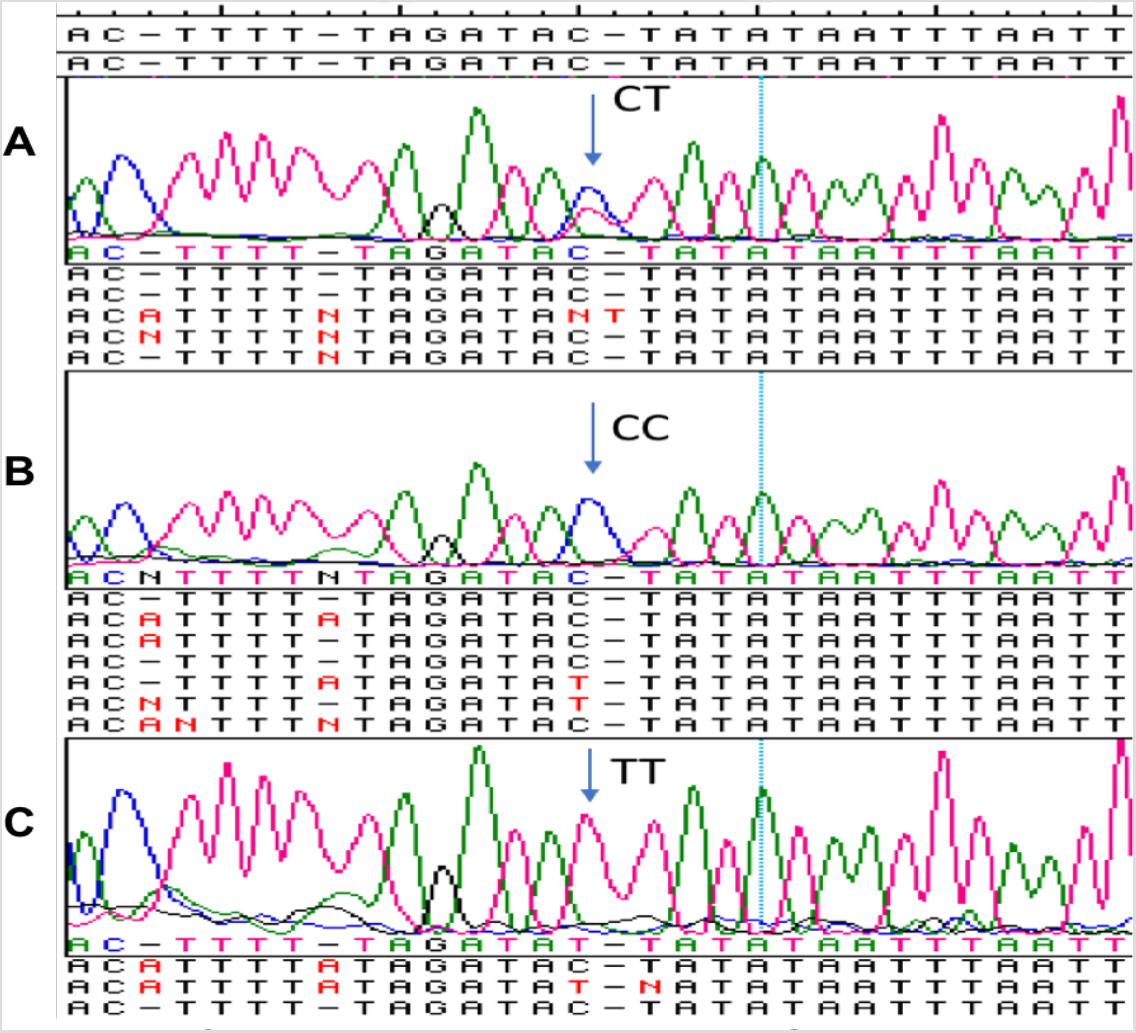
Chromatogram showing SNPs genotypes. Sequence data were visualised and aligned with reference sequence using the SeqMan pro DNASTAR software. The respective genotypes were identified at the SNP position, indicated by the arrows: (A) The CT genotype. (B) The CC genotype and (C) The TT genotype.

### Statistical Analysis

Power statistic was used to determine the minimum sample size required in order to detect at least 20% difference with 80% power at a conventional level of alpha, 0.05. Column statistics with prism 9 was used to compute percentage; mean; mean with SD (standard deviation); median; median with interquartile range and range (Table 1 and Table 3). Chi-square tests were performed to determine whether the genotypes in the populations showed deviation from Hardy-Weinberg equilibrium or not (Table 2 footnote). We computed an Odds Ratio (OR) and 95% Confidence Interval (CI) at the conventional level of alpha(0.05) with Medcalc (https://www.medcalc.org/calc/odds_ratio.php) in order to determine how strongly the presence or absence of the SNP, rs7903146 is associated with the odd of developing T2D in the population. Product Moment Correlation Coefficient Analysis was used to analyse the strength of linear relationship between two variables. SeqManPro (DNASTAR) was used to assemble the ABI trace data against the reference sequence for SNPs genotype analysis.

**Table 2:** The rs7903146 genotype and allele frequency table

| Population genotype and allele Frequency |  |  |  |  |  |  |  |  |
| --- | --- | --- | --- | --- | --- | --- | --- | --- |
|  | CC | CT | TT | C | T |  |  |  |
|  | 0.68 | 0.28 | 0.04 | 0.82 | 0.18 |  |  |  |
| Genotypes | Patients | Control | Odd Ratio (CI) | P-value | Chi-Square Test |  |  |  |
| CC | 29 | 17 | 1.71 (0.61-4.77) | 0.3086 | Genotype | CC | CT | TT |
| CT | 9 | 10 | 0.52 (0.18-1.53) | 0.2352 | Observed | 0.68 | 0.28 | 0.04 |
| TT | 2 | 1 | 1.42 (1.22-16.48) | 0.7787 | Expected | 0.67 | 0.30 | 0.03 |
| Deviation from Hardy-Weinberg Equilibrium = No |  |  |  |  | Chi-square | 0.4816 |  |  |
|  |  |  |  |  | DF | 2 |  |  |
|  |  |  |  |  | P value (two-tailed) | 0.7860 |  |  |

The genotype frequency for the genotypes, CC, CT and TT are 0.68, 0.28, and 0.04 respectively while the allele frequency for C is 0.82 and T is 0.18. The Odd ratio (Confidence Interval; CI) and P-value show that the genotypes are not associated with the odd of T2D. Computation for Hardy-Weinberg Equilibrium with Chi-Square indicated no deviation from Hardy-Weinberg equilibrium. The wild type is the CC genotype.

**Table 3:** Biochemical characteristics by genotypes in the patient and control groups

|  | **Patients** |  |  | **Controls** |  |  |
| --- | --- | --- | --- | --- | --- | --- |
|  | **CC** | **CT** | **TT** | **CC** | **CT** | **TT** |
| Age, Median IQR | 27.5 (21-41.5) | 26.5 (22-42) | 28 (20-43) | 27 (20-42) | 28 (20-42) | 27 (21-42) |
| Sex (M/F) | 13/14 | 5/4 | 1/1 | 9/8 | 5/5 | 0/1 |
| BMI, kg/m<sup>3</sup>(Median IQR) | 23.9 (21.3-26.3) | 24.0 (21.7-26.0) | 23.7.0 (22.3-26.4) | 23.6 (21.9-25.6) | 23.8 (21.8-26.1) | 23.6 (22.5-25.8) |
| FBS (mmol/L; Median IQR) | 8.40(6.4-8.58) | 9.40 (7.7-11.43)* | 10.37(7.4-11.58)* | 3.96(3.22-4.01) | 3.83(2.98-4.50) | 3.95(3.25-4.40) |
| 2-h Glucose (mmol/L), Median IQR) | 12.31(11.13-14.20) | 15.73(9.35-17.50)* | 13.90(11.73-15.50)* | 4.08(4.02-5.69) | 5.80(4.12-5.77) | 5.15(4.72-5.90) |
| C-peptide (ng/mL), Median IQR | 1.73(1.51-2.30) | 2.73(1.74-3.30)* | 2.79(1.62-3.30)* | 0.51(0.22-0.62) | 0.50(0.21-0.63) | 0.52(0.20-0.65) |
| TGs(Mean ± SD) | 2.79±1.50 | 2.82±1.40 | 3.07±1.50 | 1.46±0.59 | 1.36±1.59 | 1.92±0.54 |
| HDL (Mean ± SD) | 1.37±0.55 | 1.38±0.96 | 1.27±0.59* | 1.85±0.30 | 1.09±0.80 | 1.74±0.28 |
| LDL (Mean ± SD) | 2.89±1.26 | 3.97±1.74* | 3.93±1.64* | 1.74±0.33 | 1.54±0.41 | 1.63±0.27 |
| Total cholesterol (Mean ± SD) | 3.70±1.38 | 4.08±1.25* | 4.81±1.21* | 2.35±0.53 | 2.14±0.76 | 3.05±0.52 |
| Non-HDL (range) | 1.50-3.16 | 1.47-4.11 | 1.45-4.97 | 0.27-0.75 | 0.26-0.76 | 0.32-0.74 |

TGs = Triglycerides; SD = Standard Deviation; HDL = High Density Lipoprotein; LDL = Low Density Lipoprotein; TGs Triglycerides; FBS = Fasting Blood Sugar; BMI = Body mass index; M/F= Male/female; 2-h = 2- hour; Genotypes, CC, CT and TT. Subjects with T allele (CT and TT) in the patient group exhibits higher level of FBS, 2-h glucose; C-peptide; Total Cholesterol and elevated HDL, unique to TT genotype. Subjects with T allele in the control group showed higher blood glucose level compared to the wild type, CC, following the 2-hour glucose test. \*= statistically significantly different.

## RESULTS

The clinical demographics of the patient group is similar to that of the control group. Both sexes were represented with similar Age, height, weight, BMI, alcohol consumption, nutrition and family history (Table 1). The demographics variables in the patient group were not significantly different to the control group (Table 1). The entire population showed the GG genotype for the rs12255372 (Data not shown). However, different genotype combination, CC, CT and TT were observed with the rs7903146.

The allele frequency for rs7903146 is the study population (Fig. 2a) is lower than those reported in other part of Nigeria, the Esan, ESN and the Yoruba, YRI (Fig. 2c), though a lower population size. The total percentage of the T allele in Africa is lower compared to the Europeans and South Asia (Fig. 2b-d). The East Asian, particularly the Chinese has the lowest percentage of T- allele (Fig. 2d).

**Figure 2:**
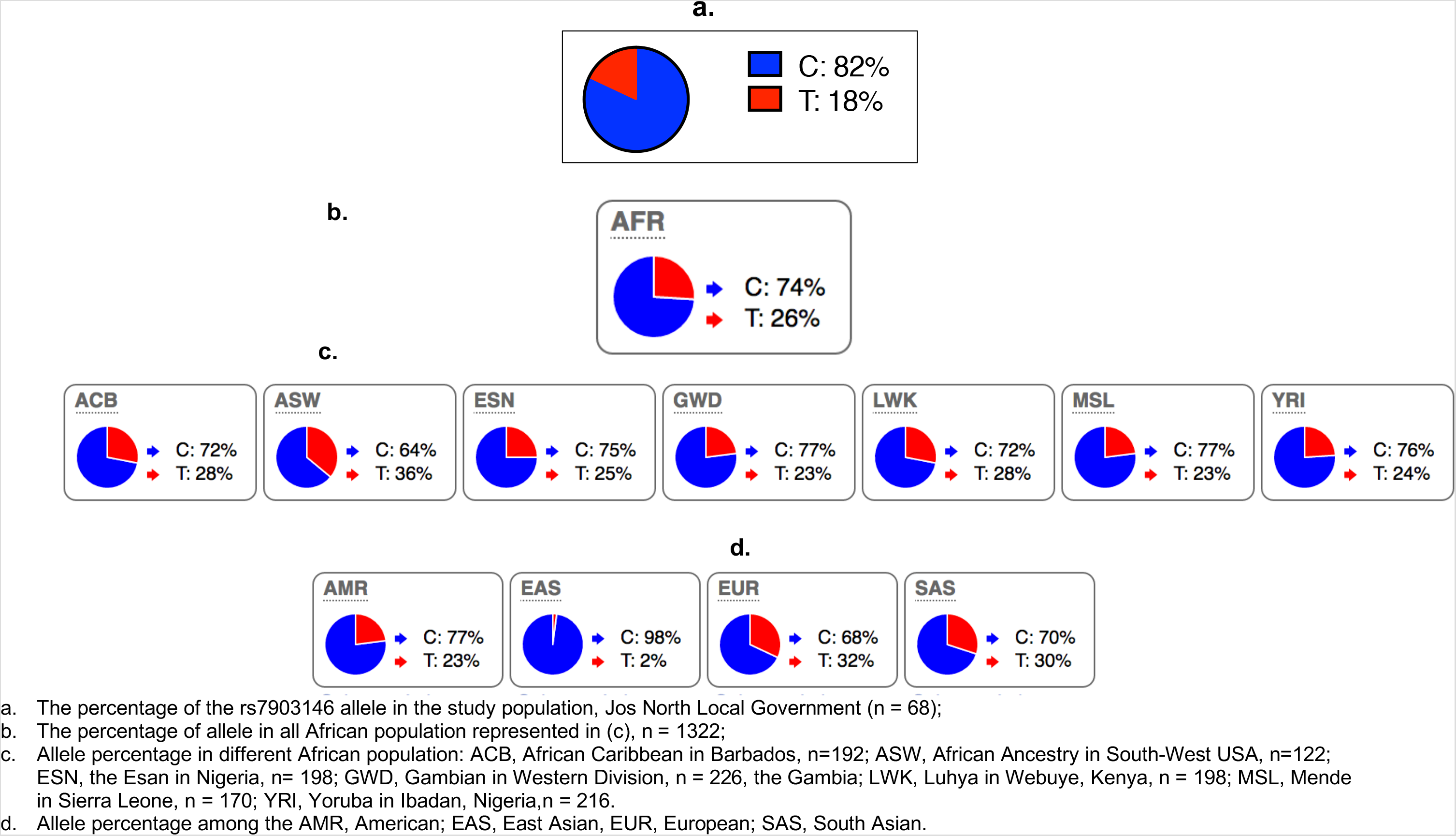
The percentage of allele in different population compared to this study population.

The genotypes, CC, CT and TT were not significantly associated with the odd of developing diabetes (Table 2). Majority of both the patient and the control groups have the wild type homozygous CC genotypes, followed by the CT, the C allele being the most common in the population (Table 2, Fig. 2). Genotypes does not show any deviation from the Hardy-Weinberg equilibrium. There is no difference in age, sex and BMI with reference to genotypes in both the patient and the control group. The two hours postprandial glucose level following the OGTT show a higher level with the CT and TT compared to the wild type. This is observed in both the patient and the control group (Table 3). The Fasting Blood Sugar (FBS) is higher in the CT and TT, the highest being in the TT genotype (Table 3). The TT shows a significantly lower HDL level in the patient group (Table 3). The LDL and total cholesterol level in the patient group is higher with CT and TT compared to the wild type, the CC (Table 3).

Product Moment Correlation Coefficient analysis (PMCC) indicates a significantly inverse correlation between C-peptide and HDL levels and a direct correlation with TGs levels (Table 4). The FBS levels inversely correlates with HDL levels but directly correlates with TGs, LDL and C-peptide levels (Table 4). Both the sugar and lipid profiles did not show any correlation with BMI (Table 4).

**Table 4:** Biochemical Parameters Correlation Analysis.

|  | **C-peptide** | **FBS** | **BMI** |
| --- | --- | --- | --- |
| **TGs** | *p =0.001,<br>r=0.491 | *p= 0.012, r=<br>0.453 | p=0.348,<br>r=0.104 |
| **HDL** | *p = 0.005, r<br>= -345 | *p= 0.006, r= -<br>0.512 | p= 0.083, r =<br>0.223 |
| **LDL** | p= 0.057, r<br>= 0.178 | *p= 0.034, r =<br>0.214 | p=0.064,<br>r=0.131 |
| **TC** | p = 0.053,<br>r= 0.055 | p= 0.074, r<br>=0.149 | p=0.062,<br>r=0.156 |
| **TC: HDL** | p = 0.084, r<br>= 0.134 | p=0.063, r=<br>0.145 | p=0.134,<br>r=0.136 |
| **C-peptide** |  | *p=0.044,<br>r=0.371 | p=0.091,<br>r=0.082 |
| **FBS** |  |  | p=0.054,<br>r=0.292 |

Product Moment Correlation Coefficient analysis (PMCC) on the biochemical parameters. TGs = Triglycerides. HDL = High Density Lipoprotein. LDL = Low Density Lipoprotein. TC = Total Cholesterol. FBS = Fasting Blood Sugar. BMI = Body Mass Index. CHOs = Carbohydrate. \*= statistically significant correlation. na = Not Applicable. N (number of patients) = 40.

## DISCUSSION

Type-2-diabetes (T2D) is of a significant global burden. The causes of T2D is multifactorial (22, 31,32). A major genetic factor is the *TCF7L2* gene, Single Nucleotide Polymorphisms (SNPs), rs7903146 (8, 22). The rs7903146 is one of the most influential variants in T2D genetic risk scores (15), thought to be modulated by obesity status, with a more significant association in the non-obese, BMI< 30kg/m^3^ (15). This SNP has been reported in different population with conflicting result. A plausible explanation to this is genetic variation and the proportion of alleles in different population (Fig. 2). As the risk of T2D is associated with the T allele, the higher the risk allele in a population, the likelihood of detecting an association with T2D.

In this study, no association was observed for the rs7903146 genotypes and the odd of T2D in the non-obese population. While this finding is in line with previous findings (33, 34), it contradicts other studies(16, 35,36). It is unclear if the lack of association is due to the low percentage of the T- allele in this study population. However, the study confirms that the level of lipids, C-peptide, and sugar in the blood could be dependent on individual rs7903146 genotypes, even without an overt association with the odd of developing T2D. This indicates that the individual SNP genotypes may be important in the management of diabetic patient.

The T allele could be associated with sub-optimal sugar metabolism: Though the blood sugar level returns to normal within two hours postprandial glucose test in all control individuals, levels in subject with the T allele is higher than levels in subjects with wild type genotype, CC. We therefore propose that the presence of this T allele among the non-obese population could be a predisposing factor to T2D, particularly in population with high carbohydrate meal, due to sub-optimal glucose metabolism.

Sugar metabolism could influence the balance of lipid profiles and vice versa, as indicated by the correlation analysis between the sugar and lipid profiles in the patient group. For example, HDL correlates negatively with both FBS and C-peptide.

Dyslipidaemia is evident in the non-obese population with diabetes. Several studies have linked cholesterol and fatty acids to insulin resistance and T2D. Elevation of Free Fatty Acids (FFAs) and TGs are key mechanisms leading to insulin resistance or impaired secretion.(37–40). A tight association is thought to exist between dyslipidaemia, insulin resistance and beta cell dysfunction (41–43).

In conclusion, this study shows that though the SNP, rs7903146 genotypes may not be overtly associated with the odd of developing T2D, both the lipid profile and blood sugar levels could be dependent on individual genotypes. Patients with T allele, are more likely to have elevated level of blood sugar, C-peptide, LDL, total cholesterol and low HDL level unlike those with the wild type. Blood sugar level in healthy subjects with the T- allele also appear to be higher than levels in subjects with the wild-type. Hence, a sub-optimal sugar metabolism is likely in subjects with the T allele and could confer the risk of developing T2D, particularly in population with heavy carbohydrate meal. These findings may be important in the management of T2D among the non-obese population.

## Data Availability

Data are available on request and following approval

## DECLARATION

### Acknowledgements

The authors thank all those that participated in the study. We thank the consultants, nurses and staff at the Faith Alive Hospital, Jos, Nigeria for their support. We thank the staffs of The National Veterinary Research Institute, Vom for an initial genetic screening. We also thank all the administrative staff at the Department of Biochemistry, university of Jos, the EBIHAS Virtual Research and Training and the British Hospital and Outpatient Clinics, Lagos, for their support for this study.

### Authors’ contributions

SS, SEA, IYL and CDL designed the study. SS and JFO carried out patient recruitment. SS and SEA carried out the laboratory experimental work. GEA, SAk, TS, SEA and SS performed the data analysis. SS, SEA, SAk, TS, JIO and GEA wrote the manuscript, which was read and approved by all authors.

